# Timing and Duration of Glucagon-like Peptide-1 Receptor Agonist Use and Risk of Nonarteritic Anterior Ischemic Optic Neuropathy

**DOI:** 10.64898/2026.02.01.26345315

**Authors:** Siar Niazi, Filip Gnesin, Baker Nawfal Jawad, Zia Niazi, Puriya Daniel Wuertz Yazdanfard, Anne P. Toft-Petersen, Kathrine Kold Sørensen, Amani Meaidi, Yousif Subhi, Christian Torp-Pedersen

## Abstract

**Purpose:** To investigate the association between glucagon-like peptide-1 receptor agonist (GLP-1RA) use and nonarteritic anterior ischaemic optic neuropathy (NAION) in type 2 diabetes, examining treatment recency and cumulative duration.

**Methods:** This nationwide registry-based nested case-control study utilised Danish health registries (1996–2023). Among 201,776 metformin-treated adults initiating second-line antihyperglycaemic therapy, 123 incident NAION cases were matched to 4,920 controls by birth year and sex (incidence-density sampling). Conditional logistic regression estimated adjusted hazard rate ratios (HRs) for GLP-1RA exposure by recency (current 0–90 days; recent 91–365 days) and cumulative duration, adjusting for socioeconomic factors, hypertension, hypercholesterolaemia, sleep apnoea, and diabetes duration.

**Results:** GLP-1RA use occurred in 63/123 cases (51.2%) and 1,688/4,920 controls (34.3%). Ever use was associated with a higher NAION rate than other second-line therapies (HR 2.13, 95% CI 1.43–3.18). Current use was associated with elevated rates (HR 2.28, 95% CI 1.49–3.48), whereas the estimate for recent use was imprecise (HR 1.69, 95% CI 0.88–3.25). By cumulative duration, no clear evidence of an increase was seen within 0–½ years (HR 0.80, 95% CI 0.32–2.05), and rates were highest at ½–1 year (HR 3.63, 95% CI 2.06–6.40) and 1–1½ years (HR 3.52, 95% CI 1.73–7.17). Findings were consistent after HbA1c adjustment and in a new-user analysis.

**Conclusion:** GLP-1RA use is associated with a higher NAION rate in type 2 diabetes. This association appears time-dependent, being most pronounced during current treatment and peaking after 6–18 months of cumulative exposure.

## Introduction

Nonarteritic anterior ischemic optic neuropathy (NAION) is the leading cause of acute optic-nerve infarction in adults over 50 years and can cause permanent visual loss (Hathaway et al. 2024, Salvetat et al. 2023). Estimated incidence is 2–10 per 100,000 persons (Foster et al. 2024, Hattenhauer et al. 1997) and it presents as sudden, painless, monocular visual loss with optic-disc swelling. No therapy has been shown to improve visual outcomes in NAION (Atkins et al. 2010, Hayreh & Zimmerman 2008). Management is therefore supportive, underscoring the need to identify modifiable risk factors to reduce risk of occurrence or recurrence.

Glucagon-like peptide-1 receptor agonists (GLP-1RAs) are increasingly prescribed for type 2 diabetes and obesity due to their glucose-lowering, weight-reducing, and cardiorenal benefits (Evans et al. 2023, Lin et al. 2021). However, a potential association with NAION has been suggested.

A single-center study reported a 4.3- to 7.6-fold higher NAION risk among semaglutide users, but referral bias limited generalizability (Hathaway et al. 2024). Subsequent epidemiological studies have reported mixed results (Abbass et al. 2025, Grauslund et al. 2024, Simonsen et al. 2025). Interpretation is constrained by NAION’s rarity, heterogeneous exposure definitions, and differences in patient characteristics and co-therapies between GLP-1RA users and nonusers. The timing of use and cumulative duration of therapy have not been examined systematically, despite their clinical relevance.

To our knowledge, no published study has jointly evaluated recency and cumulative exposure-duration categories of GLP-1RA use in relation to NAION, while addressing contemporaneous use of other second-line antihyperglycemic drugs that may influence treatment selection.

We conducted a nationwide nested case–control study in Denmark to examine the association between GLP-1RA exposure history (recency and cumulative duration) and incident NAION in adults with type 2 diabetes.

## Methods

### Study design and population

We conducted a register based, nationwide, nested case-control study investigating the association between GLP-1RA exposure and the rate of NAION.

We used the following Danish national registries: 1) The Danish National Patient Registry which contains information on all hospital admissions and outpatient contacts beginning from 1977 and up to present day (Hess 2016). Each contact is coded with a primary diagnosis and one or more secondary diagnoses according to The International Classification of Disease Eighth and Tenth Revision (ICD-8 and ICD-10) (Pedersen 2011). 2) The Danish National Prescription Registry holds information on Anatomical Therapeutic Chemical (ATC) codes, date of purchase, package size and unit size, on all prescriptions dispensed from a pharmacy since 1995 (Pottegård et al. 2017). 3) The Danish National Registry of Laboratory Results for Research contains biomarker measurements (Arendt et al. 2020). 4) The Danish Civil Registration system contains information on date of birth and gender (Pedersen 2011). 5) The Income Statistics Registry contains socioeconomic information including equivalized annual income and the Population Education Register includes the highest completed education following the International Standard Classification of Education (ISCED) (Baadsgaard & Quitzau 2011, Jensen & Rasmussen 2011). A pseudonymized version of the personal identification number assigned to all Danish citizens at birth or immigration was used to reliably link the registries.

The study population comprised individuals aged above 21 years with type 2 diabetes mellitus, identified through their first metformin prescription in the Danish National Prescription Registry. To ensure a complete medical history of diabetes treatment, only individuals whose diagnosis was confirmed on or after January 1, 1996 were included. This cutoff was chosen because the prescription registry started in 1995, allowing us to capture their first metformin prescription with at least one year of no prior use. We further restricted the population to those who initiated their first second-line antihyperglycemic therapy on or after May 9, 2007 - the date when GLP-1RA prescription was first redeemed in Denmark. Individuals with history of any type of anterior ischemic optic neuropathy (AION) before cohort entry were excluded. Eligible individuals were included from the date of initiation of second-line therapy until emigration, death, a diagnosis of giant cell arteritis, the occurrence of the outcome of interest or end of follow up (December 31, 2023). We excluded records missing education or income. The population was thus composed of Danish adults with metformin-treated type 2 diabetes on second-line antihyperglycemic treatment. Details on data sources and definitions are provided in Supplementary Table S1.

### NAION

The outcome of interest was NAION, defined using ICD-10 codes for AION (DH470C) as registered in the Danish National Patient Registry, which captures hospital-based diagnoses. To distinguish NAION from arteritic AION (AAION), we implemented a 30-day diagnostic window: all individuals with a recorded DH470C diagnosis date were followed for 30 days. If a diagnosis of giant cell arteritis (DM315 or DM316) was registered within this 30-day period, the AION event was reclassified as AAION and the patient was excluded from the case set; remaining AION cases were classified as NAION. Further information is available in Supplementary Table S1.

### Exposure

Exposure to GLP-1RAs was identified through the Danish National Prescription Registry using ATC codes A10BJ, A10AE54, A10AE56. Each redeemed prescription was assumed to cover a 60-day treatment period and linked fills less than 60 days apart as continuous use. Prescriptions gaps of more than 60 days were considered new treatment episodes. The cumulative exposure duration was obtained by aggregating for each individual up to the case index.

In addition to cumulative exposure duration, a classification based on recency of use was constructed to characterize the time elapsed since the last use of a GLP-1RA from the case index date. Individuals were categorized as current users (last exposure within 0–90 days prior to case index date) or recent users (91–365 days). Never users of GLP-1RA (users of other second-line antihyperglycemic treatment) served as the reference group.

Other second-line therapies were analogously classified and are summarized in Supplementary Table S1.

### Statistical Analysis

We provided counts and percentages for each categorical variable and tested for differences between groups using the Chi-squared test. For continuous variables, we reported the median and 1^st^ and 3^rd^ quartile. Differences between groups for continuous variables were tested using either the t-test or the Wilcoxon rank-sum test, depending on the data distribution. Each individual diagnosed with NAION was identified at the case index date and incidence density matched to forty control individuals based on sex and birth year. All analyses were done on complete cases.

Conditional logistic regression was used to estimate hazard rate ratios (HR) and 95% confidence intervals (CI) for the association between GLP-1RA use and NAION. P values <0.05 were considered statistically significant. All models were adjusted for potential confounders at case index date, including education level, income quartile, presence of hypertension, hypercholesterolemia, sleep apnea, and duration of diabetes.

To address potential confounding by glycemic control, we conducted a sensitivity analysis restricted to individuals with available HbA1c measurements during the year prior to the case index date. HbA1c was categorized using the mean HbA1c level in mmol/mol during that year into clinically relevant ranges from normal to very poor control, based on international diagnostic thresholds (Hanas, John, & International HBA1c Consensus Committee 2010). (See Supplementary Table S1).

To assess the robustness of our findings and reduce potential bias from prevalent-user inclusion, we conducted a sensitivity analysis restricted to new-users of all second-line antihyperglycemic treatments. New-users were defined as individuals whose first-ever prescription for any second-line antihyperglycemic drug was redeemed within the 365-day period preceding the case index date, with no second-line antihyperglycemic prescription prior to that window. For each case-control set, we retained the matched strata only if both the case and at least one control fulfilled this new-user criterion. Exposure was reclassified for the cumulative duration analysis of GLP-1RA treatment during the one-year window preceding the case index date, grouped into quartiles, with initiators of other second-line treatment serving as the reference category. When looking at recency of use, we distinguish current use (end of the most recent GLP-1RA treatment episode within 0–90 days before the index date) from recent use, again with new-users of other second-line treatment than GLP-1RA as reference. All models were adjusted for the same set of covariates as the primary analysis, including socioeconomic and clinical characteristics.

As a sensitivity analysis, we repeated the exposure definition using a 90-day prescription coverage period and window between consecutive prescriptions, instead of the primary 60-day definition.

The handling of data and statistics was conducted using SAS, version 9.4 (SAS Institute) and R, version 4.4.1 (R Core Team 2024) (‘R: A language and environment for statistical computing. R Foundation for Statistical Computing, Vienna, Austria. URL https://www.R-project.org/.’ n.d.).

## Results

Figure 1 outlines the study population selection process. Among 305589 adults with metformin-treated type 2 diabetes between 1996 and 2023, 233774 individuals initiated a second-line antihyperglycemic therapy on or after May 9, 2007, of whom 201776 individuals met inclusion criteria. In this cohort, 123 NAION cases were identified and incidence-density matched to 40 controls by sex and birth year, forming a study population of 5043 individuals (1,599/5,043 [31.7%] women, median age [1^st^ to 3^rd^ quartile] was 66 years [60 to 74]).

**Figure 1:**
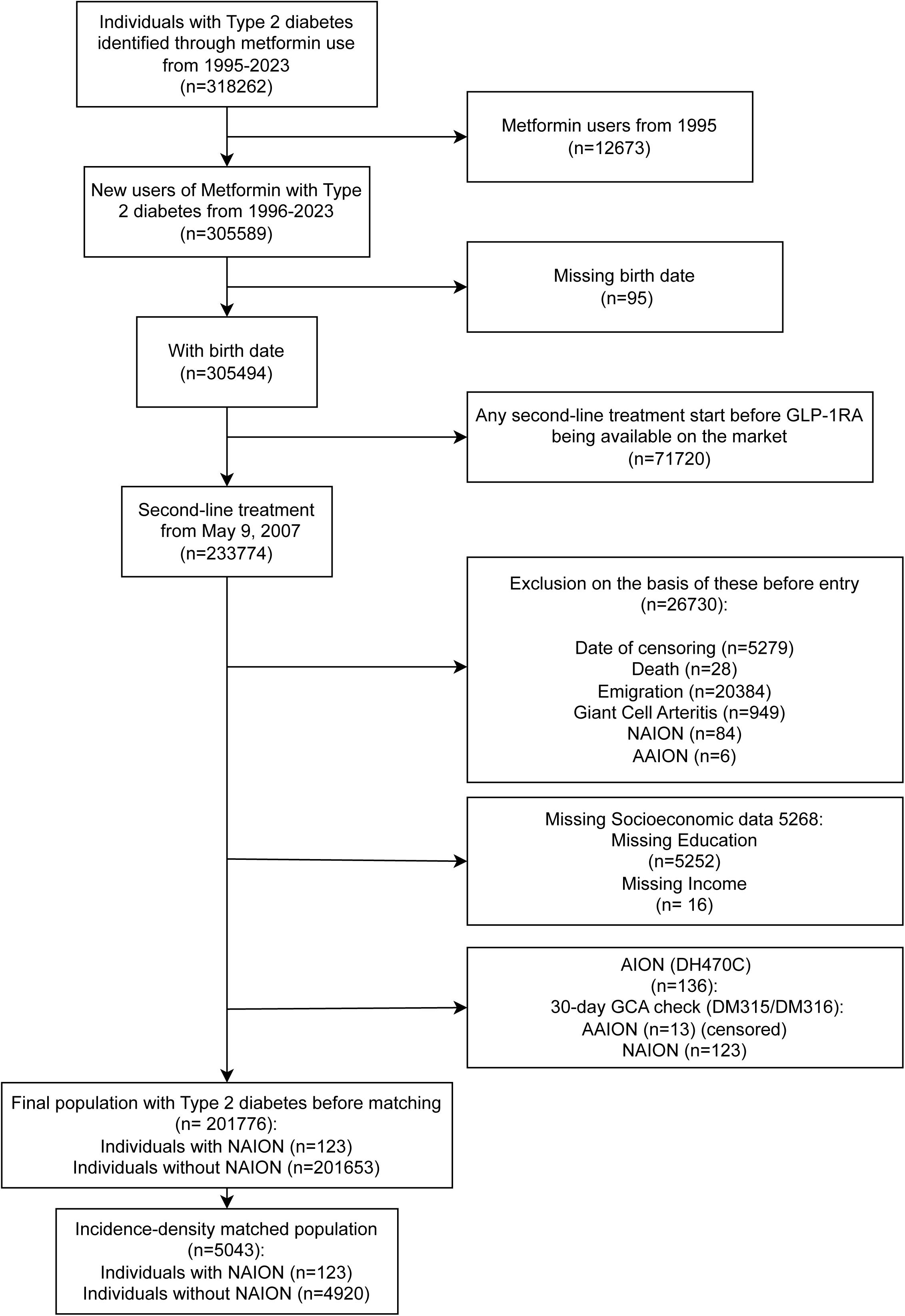
Flow Diagram of Population Selection Process. Figure 1 shows the flowchart for the cohort identification process. Nationwide Danish registry cohort of adults with type 2 diabetes identified by metformin use (1995–2023) was restricted to new metformin users with known birth date and initiation of any second-line antihyperglycemic therapy on/after 9 May 2007 (denoting the market entry of GLP-1RA). Pre-entry exclusions included censoring before eligibility (death, emigration), prior arteritic AION (AAION) or giant cell arteritis (GCA), and missing socioeconomic covariates (education or income). Final eligible population was split into individuals with NAION and without NAION; NAION cases were incidence-density matched 1:40 to controls by birth year and sex. Abbreviations: NAION, nonarteritic anterior ischemic optic neuropathy; AAION, arteritic AION; GCA, giant cell arteritis; AION, anterior ischemic optic neuropathy.

### Study Population and Baseline Characteristics

Table 1 shows characteristics measured up to the index date.

**Table 1:**
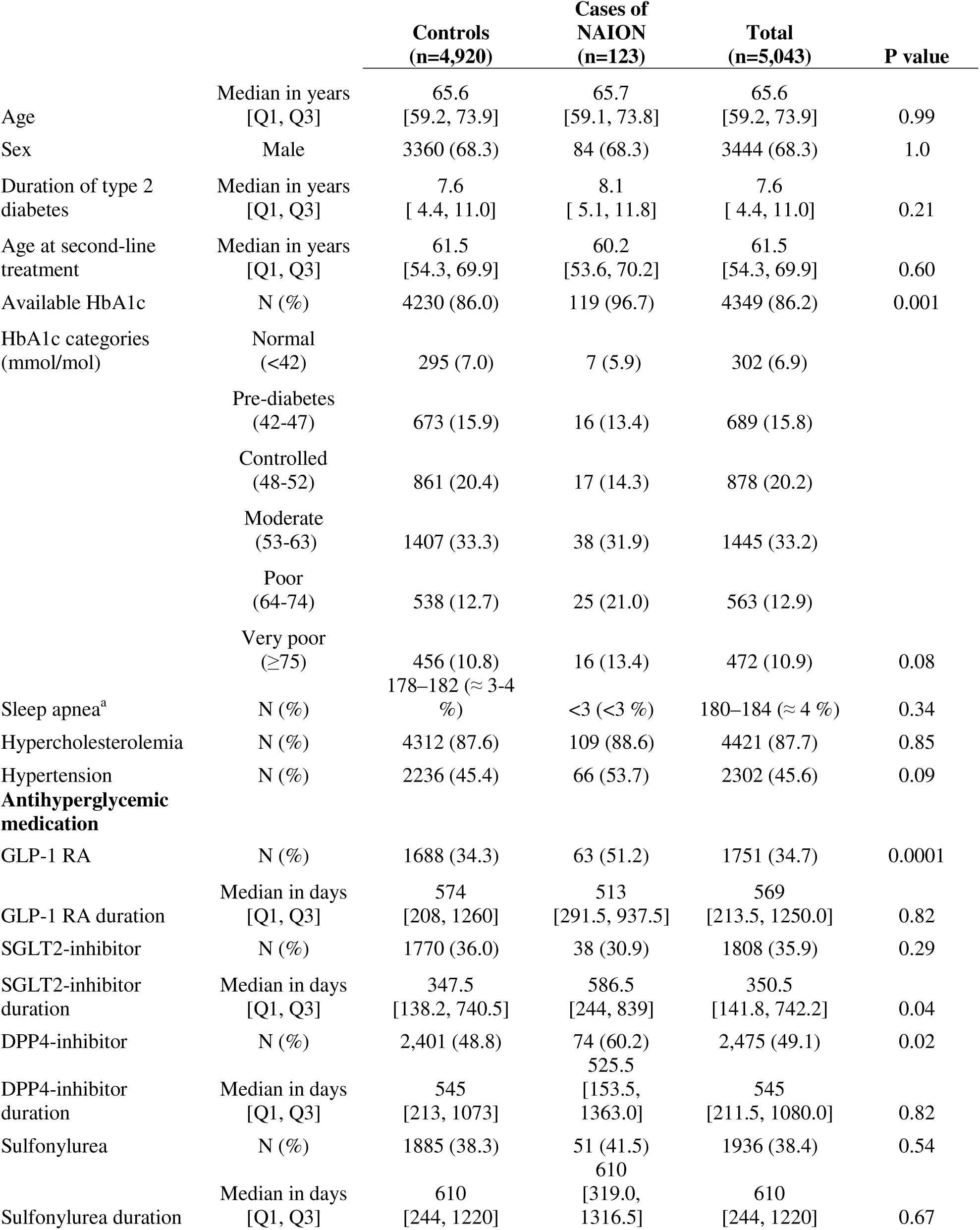
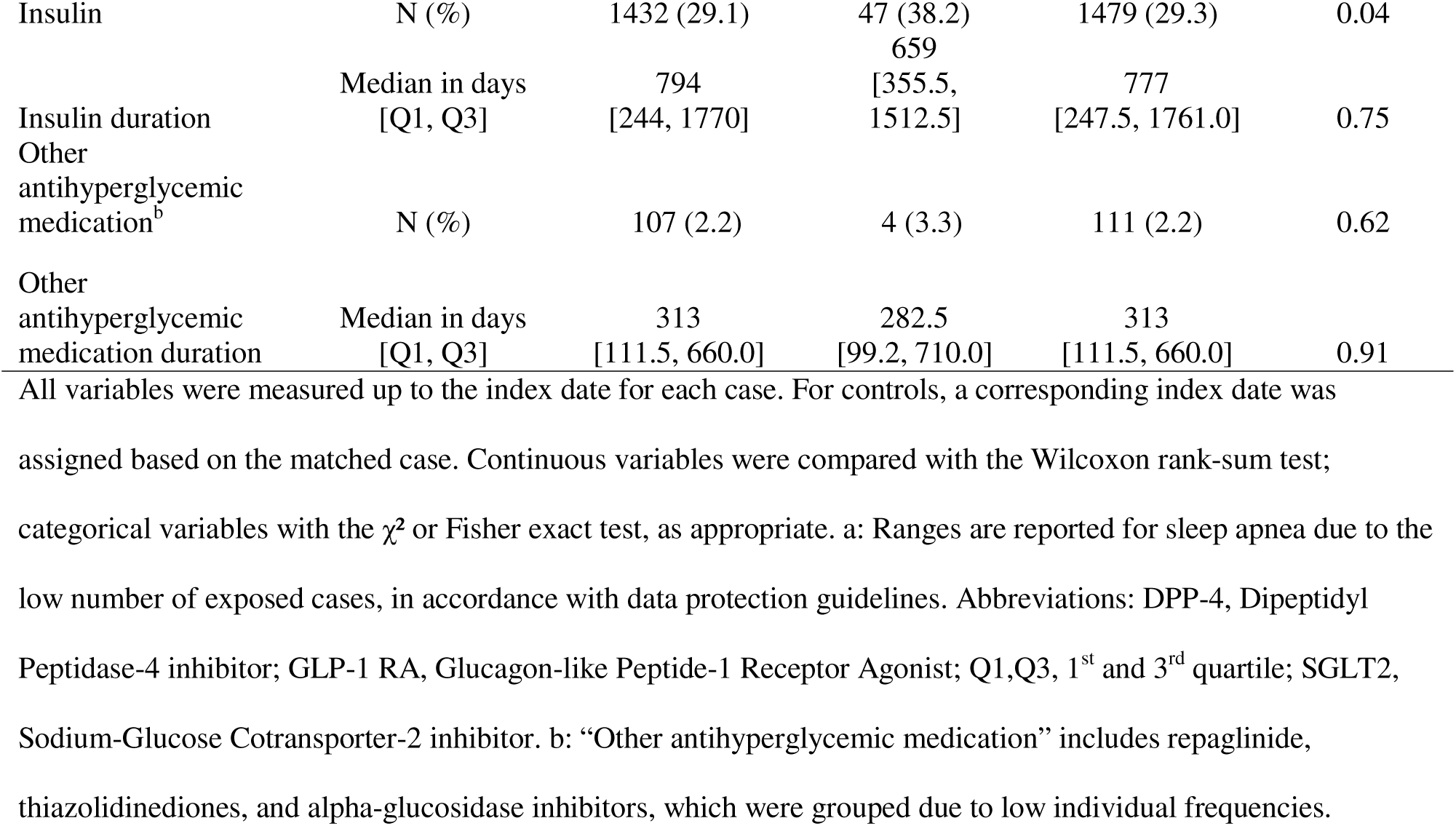
Comparative Demographics and Clinical Characteristics of Nonarteritic Anterior Ischemic Optic Neuropathy (NAION) Cases and Matched Controls at time of case index date.

There was no difference between groups in duration of type 2 diabetes (8.1 years in cases versus 7.6 years in controls; P=0.21), nor age at second-line therapy initiation (60.2 years versus 61.5 years; P=0.60).

Cases were more likely to have had a recorded HbA1c value within one year prior to case index date (96.7% versus 86.0%; P<0.001). The distribution across HbA1c severity bands did not differ significantly between cases and controls (P=0.08).

The prevalence of sleep apnea, hypercholesterolemia, and hypertension was similar between cases and controls. Use of second-line antihyperglycemic agents varied modestly between groups. Cases were more likely to have received a GLP-1RA (51.2% versus 34.3%; P<0.001). The median cumulative duration of GLP-1RA exposure did not differ between cases and controls (513 versus 574 days; P=0.82). Similarly, cases were more likely to have used dipeptidyl peptidase-4 (DPP-4) inhibitors (60.2% versus 48.8%; P=0.02) and insulin (38.2% versus 29.1%; P=0.04). Sodium-Glucose Co-Transporter-2 (SGLT2) Inhibitor use was not more prevalent among cases (30.9% versus 36.0%; P=0.29), but cumulative exposure duration in days was longer (median 587 versus 347; P=0.04). Other drug classes, including sulfonylureas, showed no meaningful differences.

### Primary Analysis

Figure 2 shows the association between GLP-1RA use and NAION. GLP-1RA use was associated with a significantly higher rate of NAION compared to never users of GLP-1RA, with a HR of 2.13 (95% CI, 1.43 to 3.18, P<0.001).

**Figure 2:**
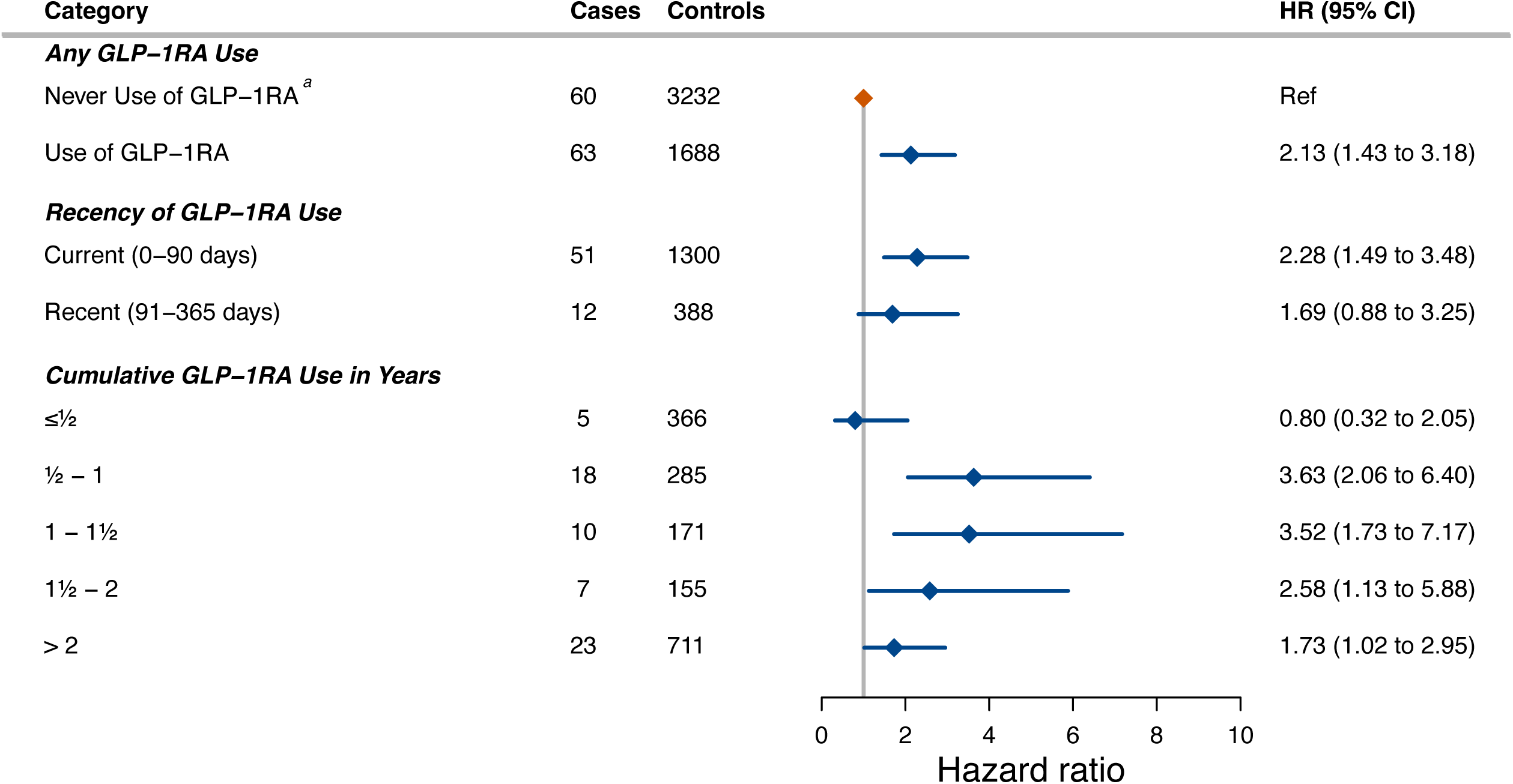
Association Between GLP-1 Receptor Agonist Use and Risk of NAION. Figure 2 shows a forest plot of adjusted hazard rate ratios (HRs) with 95% CIs for NAION comparing categories of GLP-1RA exposure among metformin-treated adults initiating second-line therapy in three separate conditional logistic regression models. In all three models, never-users of GLP-1RA were used as the reference group. In the first model, any GLP-1RA use was compared against never use, in the second model, GLP-1RA exposure was subdivided into groups of recency of use (current ≤90 days; recent 91–365 days), and in the third model, GLP-1RA exposure was investigated according to cumulative GLP-1RA duration (0–½, ½–1, 1–1½, 1½–2, >2 years). a: reference category. Abbreviations: GLP-1RA, glucagon-like peptide-1 receptor agonist; NAION, nonarteritic anterior ischemic optic neuropathy; HR, hazard rate ratio; CI, confidence interval.

Current users had a significantly higher rate of NAION than never users of GLP-1RA (HR, 2.28; 95% CI, 1.49 to 3.48, P<0.001). No significant association was observed for users who recently stopped, although the point estimate showed a higher rate.

When categorized by cumulative GLP-1RA exposure duration, a duration response pattern was seen. Individuals with 0–½ years of GLP-1RA treatment did not show clear evidence of an increased rate (HR, 0.80; 95% CI, 0.32 to 2.05, P=0.65) compared to never-users of GLP-1RA. The highest rate was observed in the ½–1-year (HR, 3.63; 95% CI, 2.06 to 6.40, P<0.001) and 1–1½ years (HR, 3.52; 95% CI, 1.73 to 7.17, P<0.001) groups compared to never-users of GLP-1RA. Rates were attenuated but remained elevated for 1½–2 years (HR, 2.58; 95% CI, 1.13 to 5.88, P=0.02) and for those with over two years of cumulative exposure (HR, 1.73; 95% CI, 1.02 to 2.95, P=0.04).

### New-user subgroup

Figure 3 shows the subgroup analysis restricted to new users only, defined as individuals initiating their first-ever second-line prescription within a year prior to the case index date. In the new-user analysis, GLP-1RA initiators (17 cases, 363 controls) had a higher NAION rate than initiators of other second-line therapy (20 cases, 980 controls) (HR 2.46, 95% CI, 1.09 to 5.57; P=0.03).

**Figure 3:**
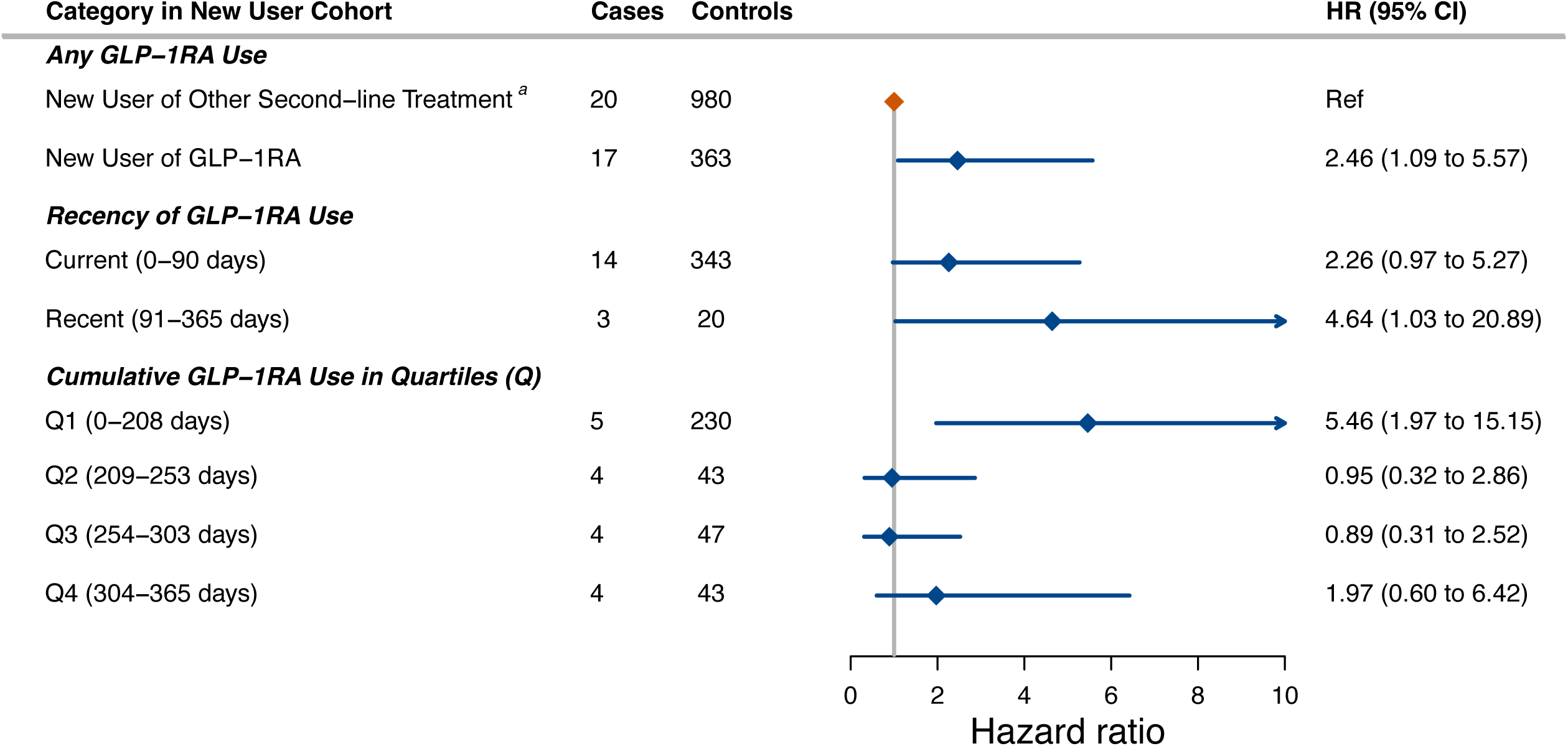
Association Between GLP-1 Receptor Agonist Use and Risk of NAION in a New-user Subgroup. Figure 3 shows forest plot restricted to the new-user cohort of second-line therapies. Comparisons include any new use of GLP-1RA versus new use of other second-line treatment, recency of GLP-1RA use (current ≤90 days; recent 91–365 days), and cumulative GLP-1RA use in quartiles over the first year (Q1: 0–208 days; Q2: 209–253; Q3: 254–303; Q4: 304–365). Points are HRs with 95% CIs from conditional logistic regression matched on sex and birth year and adjusted as in the main model. a: reference category. Abbreviations: GLP-1RA, glucagon-like peptide-1 receptor agonist; NAION, nonarteritic anterior ischemic optic neuropathy; HR, hazard rate ratio; CI, confidence interval; Q, quartile.

Consistent with the full study population, current use showed a trend of elevated NAION rate (HR, 2.26; 95% CI, 0.97 to 5.27, P=0.06), while users of GLP-1RA who recently stopped, were associated with higher risk estimates but with high uncertainty (HR, 4.64; 95% CI, 1.03 to 20.89, P=0.05), reflecting sparse data (only three exposed cases).

Stratification of cumulative exposure by quartiles in this group revealed the highest risk estimate was in the 1^st^ quartile (0-208 days; HR 5.46; 95% CI 1.97 to 15.15, P=0.001).

### HbA1c-adjustment and sensitivity analyses

To address potential confounding by glycemic control, analysis was repeated in the subgroup with available HbA1c data (119 cases, 4,230 controls) and was adjusted for HbA1c levels. In this subgroup, use of GLP-1RA remained significantly associated with a higher NAION rate (HR, 2.09; 95% CI, 1.39 to 3.14, P<0.001). (See Supplementary Figure S1).

Adjusting for SGLT2, DPP-4, and insulin showed HR 2.16 (95% CI, 1.43 to 3.24; P<0.001). Defining exposure with 90-day coverage/windows produced similar results (See Supplementary Figure S1).

## Discussion

In this nationwide, registry-based nested case-control study of Danish adults with metformin-treated type 2 diabetes receiving second-line antihyperglycemic treatment, we found that current use of GLP-1RAs was associated with a higher rate of NAION compared to users of other second-line treatments. Current use of GLP-1RAs was associated with an approximate two-fold increase in the rate of NAION. The excess rate was most pronounced during the first 6–18 months of cumulative exposure. Longer-term exposure (> 2 years) still conferred some evidence for the association. The association was not evident among individuals with less than six months of cumulative GLP-1RA exposure (HR 0.80, 95% CI 0.32 to 2.05).

However, estimates in this early-duration category were imprecise due to sparse exposed events, and thus do not in isolation establish a minimum treatment duration threshold.

### Adjustment and Robustness of Findings

Cases and controls were matched for age and sex and had comparable age at second-line treatment and diabetes duration. After adjusting for income, education, sleep apnea, hypercholesterolemia, hypertension, and diabetes duration, findings persisted and were robust to exclusion of prevalent users. The association persisted after adjusting for recent HbA1c levels, arguing against confounding by glycemic control or survivorship.

The clearest divergence between cases and controls lies in pharmacologic complexity. Cases were more likely to receive GLP-1RAs (51.2 % vs 34.3 %), DPP-4 inhibitors (60.2 % vs 48.8 %), insulin (38.2 % vs 29.1 %), and accumulated more exposure to SGLT2 inhibitors. After adjusting in the sensitivity analyses, the differences in pharmacotherapy did not change the association, supporting the robustness of the primary findings.

### Comparison With Previous Studies

Previous studies have used the Danish registries to investigate the association between GLP-1RAs and NAION, also finding similar associations, although with different population and with crudely defined exposure and outcome definitions (Grauslund et al. 2024, Simonsen et al. 2025). Our study provides novel and crucial knowledge on the recency and the duration of treatment while using an outcome definition which closer resembles clinical practice.

### Biological Plausibility

NAION is believed to arise from transient hypoperfusion of the optic-nerve head supplied by the short posterior ciliary arteries. A small, crowded optic disc with little cup, confers structural vulnerability. Known systemic risk factors are vascular comorbidities such as hypertension, diabetes, dyslipidemia, and nocturnal hypotension, as well as obstructive sleep apnea, smoking and vasoactive medications, but the mechanistic evidence remains incomplete (Evans et al. 2023, Lin et al. 2021, Salvetat et al. 2023).

Although the mechanistic underpinnings remain speculative, several biological pathways may be relevant. GLP-1 receptors are expressed on vascular smooth-muscle and endothelial cells stimulating vasodilation and natriuresis, lowering daytime and nocturnal blood pressure (Chai et al. 2012, Cigrovski Berkovic & Strollo 2023). In predisposed individuals, these shifts could compromise optic-nerve head perfusion.

GLP-1RAs achieve more rapid glycemic reduction than most other antihyperglycemic drugs (Garber et al. 2009, Nauck et al. 2009, Pratley et al. 2010). Swift blood sugar drops have been linked to early worsening of retinal microangiopathy in trials such as SUSTAIN-6 (Marso et al. 2016). Rapid osmotic equilibration after such declines could trigger fluid shifts and swelling, while natriuresis which intensifies nocturnal hypotension may provoke ischaemia (Vujosevic et al. 2025).

Diabetic papillopathy is a self-limiting condition with optic-disc oedema linked to diabetic microangiopathy, often with good visual outcome, but in some cases may cause irreversible damage (Aguiar et al. 2024, Giuliari et al. 2011, Ostri et al. 2010). In some rare cases, diabetic papillopathy has been later diagnosed as a frank NAION. The overlap in vascular dysregulation raises the possibility that haemodynamic changes from GLP-1RA use might affect this conversion. GLP-1RAs signaling modulated retinal microglia, and dysregulated neuro-inflammation in preclinical studies (Puddu & Maggi 2022, Zhou et al. 2021). Microglia and macrophage-driven inflammation is implicated in secondary injury after optic-nerve ischemia in primates. Thus, dysregulated neuro-inflammation could plausibly impede recovery after transient hypoperfusion (Salgado et al. 2011). Further investigation is needed to clarify this potential mechanism.

### Clinical implications

Across analyses, findings support heightened vigilance for NAION during current GLP-1RA treatment, with estimates appearing most pronounced within 6–18 months of cumulative exposure. Sudden painless vision loss or new visual-field defects always warrant urgent assessment regardless of treatment status. These results support proactively reinforcing this advice in patients with type 2 diabetes receiving GLP-1RAs during this window.

### Generalizability

Denmark’s universal healthcare and pharmacy-dispensing registries minimize selection bias, yet prescribing thresholds and comorbidity spectra differ in countries where GLP-1RAs are reserved for obesity or cardiovascular indications. Caution is therefore warranted in generalizing these findings.

### Study Strength and Limitations

Strengths of this study lie in its study design, including nationwide, complete capture of drug dispensations and hospital diagnoses.

Due to NAION’s rarity, many of our subgroup analyses showed numerically elevated point estimates with a wide confidence interval reflecting sparse data. NAION cases managed in primary care may be missed, as they are not referred to hospitals and therefore not captured in the registries. Nevertheless, we do not expect hospital referral to differ by GLP-1RA use among individuals with type 2 diabetes, and thus do not anticipate selection that would otherwise have impacted our findings.

HbA1c is not recorded uniformly across individuals, and differential measurement could reflect clinical follow-up intensity. However, NAION typically presents as acute, symptomatic vision loss that prompts urgent evaluation, making it less likely that laboratory measurement patterns determined outcome ascertainment. While surveillance bias cannot be excluded, findings were consistent in the subgroup of individuals with HbA1c measurements.

We were able to control for multiple confounding factors including hypertension, hypercholesterolemia, sleep apnea, and duration of diabetes. However, residual confounding remains possible due to missing data on body mass index (BMI) and smoking status. In Denmark, smoking status and BMI are highly correlated with education and income, which we adjusted for, so part of this lifestyle-related confounding is likely captured (National Institute of Public Health 2022).

## Conclusion

Use of GLP-1RA was associated with an increased rate of NAION in adults with type 2 diabetes, particularly among current users and those with 6–18 months of cumulative exposure. Clinicians prescribing GLP-1RAs should monitor patients for early symptoms of optic nerve ischemia such as sudden blurred vision, visual field defects or color desaturation. Prospective and mechanistic studies are warranted to assess causality and identify high-risk subgroups.

## Data Availability

Individual-level data are available only through authorization from Statistics Denmark and the Danish Health Data Authority and cannot be shared publicly due to data protection regulations. Access may be obtained by application to Statistics Denmark, subject to relevant approvals. Study protocol and analysis code are available from the corresponding author on reasonable request.

## Ethics

This project was approved by the Knowledge Centre on Data Protection Compliance, The Capital Region of Denmark. In Denmark, informed consent or review by a research ethics committee is not required for register based studies. The study adhered to the Declaration of Helsinki and STROBE (von Elm et al. 2008, World Medical Association 2013).

## Funding

The study was supported by a Research Grant from North Zealand Hospital (grant: E-20227-40-31). The funding organization had no role in the design or conduct of this research.

## Transparency declaration

The lead author (SN) affirms that this manuscript is an honest, accurate, and transparent account of the study being reported; that no important aspects of the study have been omitted; and that any discrepancies from the study as planned have been explained.

## Contributors

SN and CTP conceived and designed the study. SN acquired the data. SN, CTP, AM, and YS analysed the data. All authors contributed to interpretation of the data. SN drafted the manuscript, and all authors critically revised it for important intellectual content and approved the final version. SN is the guarantor and accepts full responsibility for the work and the conduct of the study, had access to the data, and controlled the decision to publish.

## Notes

### Competing Interest Statement

The authors have declared no competing interest.

### Author Declarations

Knowledge Centre on Data Protection Compliance, The Capital Region of Denmark

