## Supplementary Material for "Timing and Duration of Glucagon-like Peptide-1 Receptor Agonist Use and Risk of Nonarteritic Anterior Ischemic Optic Neuropathy"

*Supplementary Figure S1: Subgroup Analyses in Patients with HbA1c Measurements and Adjustments for SGLT-2 inhibitors, DPP4 inhibitors and Insulin*

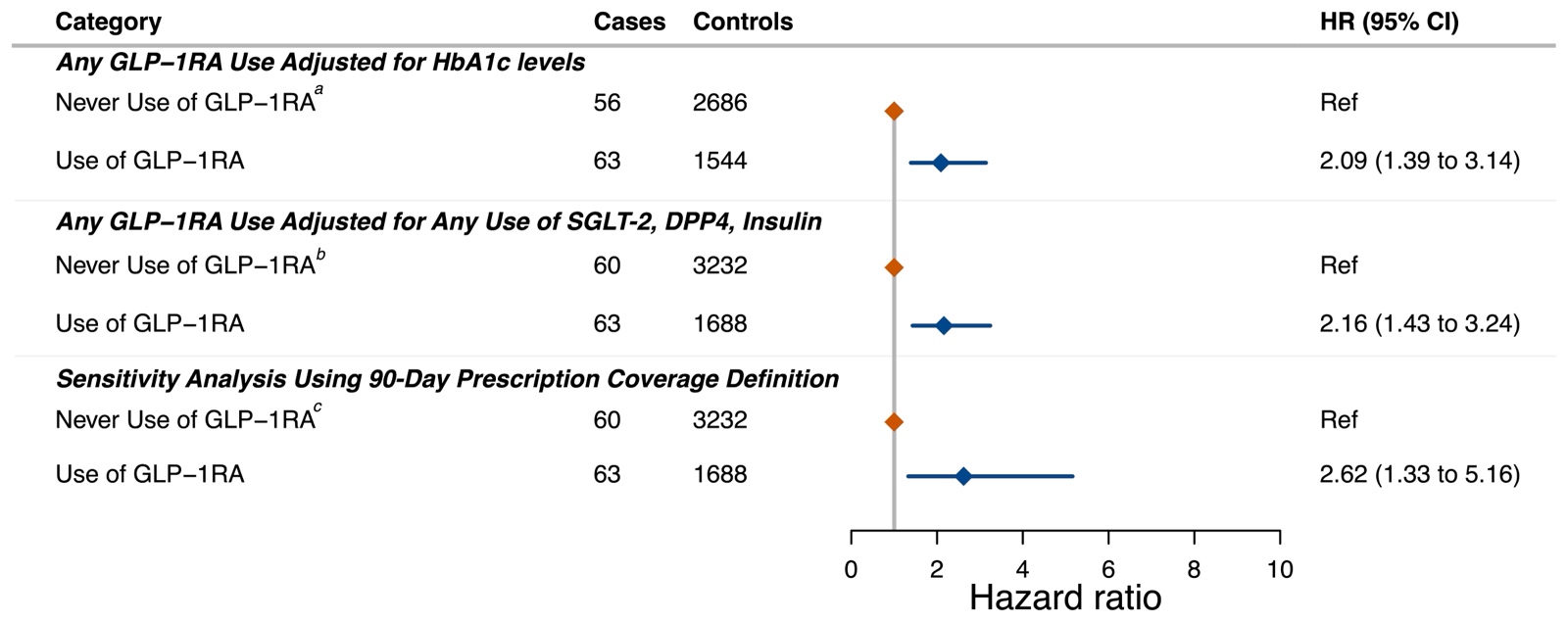

Supplementary Figure S1 shows three separate models with adjusted HRs (95% CIs) for any GLP-1RA use versus never use. In the 1^st^ analysis was adjusted for HbA1c measurements where those were available. In the 2^nd^ analysis the primary model was adjusted for use of SGLT-2 inhibitors use, DPP-4 inhibitors use, or insulin use. The 3^rd^ model was a sensitivity analysis defining exposure to medication as 90 days, instead of 60 days as in the main analysis. All models were matched on sex and birth year and adjusted for hypertension, hypercholesterolemia, sleep apnea, education, income and duration of diabetes. a, b, c: were used as the reference group, who were users of other second-line antihyperglycemic therapies with no prior GLP-1RA exposure. Abbreviations: HbA1c, hemoglobin A1c; SGLT-2, sodium–glucose co-transporter-2; DPP-4, dipeptidyl peptidase-4; HR, hazard rate ratio; CI, confidence interval; GLP-1RA, glucagon-like peptide-1 receptor agonist; NAION, non-arteritic anterior ischemic optic neuropathy.

*Supplementary Table S1: Data Sources and Data Definitions*

| **Variable** | **Registry** | **Data availability** | **Classification** | **Definition** | | |
| --- | --- | --- | --- | --- | --- | --- |
| **Medication** | | | | | | |
| Antihyperglycemics | The National Prescription Registry | 1995-2023 | The Anatomical Therapeutic Chemical Classification System (ATC) | Insulin | A10A | |
|  |  |  |  | Metformin (Biguanide) | A10BA02, A10BD02 - A10BD27 | |
|  |  |  |  | Sulfonylurea | A10BB - A10BD06 | |
|  |  |  |  | Alpha-Glucosidase | A10BF, A10BD17 | |
|  |  |  |  | Thiazolidinedione | A10BG, A10BD03 - A10BD12, A10BD26 | |
|  |  |  |  | Dipeptidyl Peptidase IV (DPP4) Inhibitors | A10BH, A10BD07 - A10BD13, A10BD18 - A10BD19, A10BD21 - A10BD22, A10BD24 - A10BD25, A10BD27 | |
|  |  |  |  | Sodium-Glucose Co-Transport Protein 2 (SGLT2) Inhibitors | A10BK, A10BD15 - A10BD16, A10BD19 - A10BD21, A10BD23 - A10BD25, A10BD27 | |
|  |  |  |  | Glucagon-like peptide-1(GLP-1) receptor agonist | A10BJ, A10AE54, A10AE56 | |
|  |  |  |  | Repaglinide | A10BX02, A10BD14 | |
| **Diagnoses** | | | | | | |
| Anterior Ischemic Optic Neuropathy  (AION) | The Danish National Patient Register | 1977-2023 | The International Classification of Diseases and Related Health Problems, 10th & 8th revision (ICD-8 & ICD-10) | DH470C | | |
| Hypertension | The Danish National Patient Register |  | (ICD-8 & ICD-10)  &  The Nordic Medico-Statistical Committee Classification of Surgical Procedures (Nomesco, NCSP) | 401-404, 41009, 41099, DI10, DI109, DI11, DI110, DI119, DI119A, DI12-DI129, DI129A, DI13-DI139, DI15-DI159 | | |
| Sleep Apnea | The Danish National Patient Register |  | (ICD-8 & ICD-10) | DG4732 | | |
| Giant Cell Arteritis | The Danish National Patient Register |  | (ICD-8 & ICD-10) | DM315, DM316 | | |
| Hypercholesterolemia | The Danish National Patient Register |  | (ICD-8 & ICD-10)  &  (ATC) | Familial | DE780B, DE780B1  DE780B2 | |
|  |  |  |  | General | DE780A, DE780C, DE780D, DE780E, DE781, DE782, DE783, DE784, DE785 | |
|  | The National Prescription Registry |  |  | C10AA, C10AB, C10BX, C10BA01, C10BA02, C10BA04, C10BA05, C10BA06, C10BA10, C10AX09, C10BA03, C10BA09, C10BA08, C10BA07, C10AX12, C10AX13, C10AX14, C10AC01, C10AC02, C10AC03, C10AC04 | | |
| **Blood Biomarker** | | | | | | |
| Hemoglobin A1c (HbA1c) | The Danish National Registry of Laboratory Results for Research | 2008-2023 | Nomenclature for Properties and Units (NPU) classification | Levels in mmol/mol  Normal: (<42)  Pre-diabetes: (42-47)  Controlled: (48-52)  Moderate: (53-63)  Poor: (64-74)  Very poor (≥75) | | NPU27300, NPU29296, NPU03835, NPU02307 |
| **Measured Socio-economic Confounding Factors** | | | | | | |
| Education | The Danish Education Registry | 1980-2023 | Highest obtained education - International Standard Classification of Education (ISCED) | Elementary school only; secondary school only; skilled worker; theoretical education; theoretical and research education | | |
| Income | The Income Statistics Registry | 1970-2023 | equivalized annual income divided according to quartiles | Low income (below first quartile); Middle income (first-third quartile); High income (above third quartile) | | |

Supplementary Table S1 lists the registries and classification systems used for the identification of medications, diagnoses, blood biomarkers, and socio-economic confounders in the nationwide nested case-control study. Medication data were obtained from the Danish National Prescription Registry and classified according to the Anatomical Therapeutic Chemical (ATC) system. Diagnoses of anterior ischemic optic neuropathy (AION), hypertension, sleep apnea, giant cell arteritis, and hypercholesterolemia were extracted from the Danish National Patient Register using International Classification of Diseases (ICD-8 & ICD-10) and Nordic Medico-Statistical Committee (NOMESCO) procedure codes; hypercholesterolemia was also captured using ATC codes. HbA1c values were retrieved from the Danish National Registry of Laboratory Results for Research and classified according to the Nomenclature for Properties and Units (NPU) system. Socio-economic variables (education and income) were derived from the Danish Education Registry and the Income Statistics Registry and categorized using international standards and equivalized annual income in quartiles, respectively.
